# COVID-19 pandemic in the African continent: Forecasts of cumulative cases, new infections, and mortality

**DOI:** 10.1101/2020.04.09.20059154

**Authors:** T Achoki, U Alam, L Were, T Gebremedhin, F Senkubuge, A Lesego, S Liu, R Wamai, Y Kinfu

## Abstract

**Background:** The epidemiology of COVID-19 remains speculative in Africa. To the best of our knowledge, no study, using robust methodology provides its trajectory for the region or accounts for local context. This paper is the first systematic attempt to provide prevalence, incidence, and mortality estimates across Africa.

**Methods:** Caseloads and incidence forecasts are from a co-variate-based instrumental variable regression model. Fatality rates from Italy and China were applied to generate mortality estimates after making relevant health system and population-level characteristics related adjustments between each of the African countries.

**Results:** By June 30 2020, around 16.3 million people in Africa will contract COVID-19 (95% CI 718,403 to 98,358,799). Northern and Eastern Africa will be the most and least affected areas. Cumulative cases by June 30 are expected to reach around 2.9 million (95% CI 465,028 to 18,286,358) in Southern Africa, 2.8 million (95% CI 517,489 to 15,056,314) in Western Africa, and 1.2 million (95% CI 229,111 to 6,138,692) in Central Africa. Incidence for the month of April 2020 is expected to be highest in Djibouti, 32.8 per 1000 (95% CI 6.25 to 171.77), while Morocco will experience among the highest fatalities (1,045 deaths, 95% CI 167 to 6,547).

**Conclusion:** Less urbanized countries with low levels of socio-economic development (hence least connected to the world), are likely to register lower and slower transmissions at the early stages of an epidemic. However, the same enabling factors that worked for their benefit can hinder interventions that have lessened the impact of COVID-19 elsewhere.

## I. INTRODUCTION

The novel human coronavirus (SARS-CoV-2 or COVID-19) outbreak initially emerged in Wuhan, China, in late 2019. Since then, it has spread to 198 countries and territories around the world and has been declared a pandemic. (1) As of March 31, 2020, there have been 750,874 reported COVID-19 cases, with 36,045 deaths, and 117,603 recoveries. (1) The first confirmed COVID-19 case in Africa was reported in Egypt on February 14, 2020 and since then the number of confirmed infections in the region has surpassed 5,000 cases as of March 31, 2020. (2-3) Comoros, Lesotho, Malawi, and South Sudan are the only African countries that have not reported a confirmed case as of March 31, 2020.

To date, no vaccine or effective treatment is available for COVID-19. Therefore, the ability to minimise the devastating consequences of the disease on people’s lives and livelihoods relies on the implementation of effective preventative non-pharmaceutical interventions (NPIs). NPIs include multiple public health measures designed to reduce viral transmission rates in a population by reducing the reproduction number (R_0_); the average number of secondary cases each case generates. (4-6)

NPIs directly influence the course of the COVID-19 pandemic, including the rate of spread, and the expected duration of the pandemic. However, several characteristics of the virus remain ambiguous or mostly unknown, (7-8) such as the incubation period (the time between infection and symptom onset), serial interval (the time between symptom onset of a primary and secondary case) the extent of asymptomatic cases, the possibility of pre-symptomatic infectiousness, case fatality rate (CFR), and also the possible role of weather in transmission. Estimates for COVID-19 CFR range from 0.3-1%. (9) Asymptomatic or mild presentation comprises the bulk of the reported cases, which is an estimated at 80%. (10) Longitudinal viremia measurements from a small study (sample size of 16), suggests that there are high enough viral loads to trigger pre-symptomatic infectiousness for 1-2 days before the onset of symptoms.(11) With the lack of clinical studies measuring viremia, the infectious period also remains largely unknown, with estimates ranging from few days to 10 days or more after the incubation period. (11)

The complexity of the infection and recovery process, therefore, means that proper understanding of the epidemiological dynamics of COVID-19 within the local context is fundamental to combat the pandemic. Studies illustrating future trajectories of the disease are not only helpful to develop early warning systems, avoid overwhelming healthcare services and minimize morbidity and mortality from the disease, but also assist countries to evaluate the effects of interventions and the long-term consequences of the virus on peoples’ livelihood. This is particularly true in Africa, where livelihoods are fragile, and previous epidemics, such as HIV/AIDS and, more recently, Ebola, have been known to exert enormous socioeconomic consequences. (12-14) In addition, in the face of a new pandemic in the region, the already overstretched healthcare systems that are struggling to deliver essential healthcare services such as immunization and HIV/AIDS treatment would be in further jeopardy and at risk of losing the gains achieved so far in disease control efforts. This study, using a robust methodology, provides spatial and temporal trajectories of COVID-19 for the entire Africa region. To the best of our knowledge, this is the first such attempt and accounts for the local context and characteristics relevant to the epidemiology of the diseases in the region.

The remainder of this paper is divided into three sections. In section 2, we describe our covariate based predictive model and the input data that has been used in the modeling exercise. We also describe the methodology used for estimating deaths from COVID-19. Section 3 provides the results on the projected new infections, cumulative cases, and deaths due to COVID-19 across several countries, covering the five sub regions. Finally, we discuss the key findings and implications of the study in section 4 and in Section 5 provide some concluding remarks.

## II. METHODS

The analysis is based on a linear instrumental variable (IV) regression framework. The basic model consists of two equations which can be expressed as follows: (15-17)

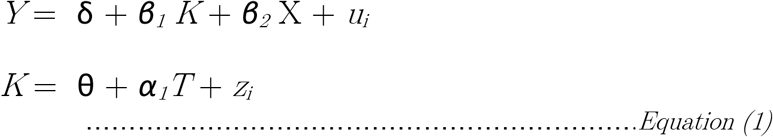

Here, *Y* represents the number of confirmed cases of COVD-19 for each country as of March 31, 2020. *K* is the rate of infection between week 1 and week 2 of the epidemic in the country and is an endogenous variable. *T* and *X* represent a set of instrumental and exogenous variables, respectively. *u*_*i*_ and *z*_*i*_ are error terms and both are assumed to have a zero mean and a nonzero-correlation. (18-20) δ, θ, *α*, *β* are regression coefficients to be estimated from the model.

The model captures a combination of epidemiological, socio-economic and health system readiness related covariates. On the epidemiological side, it includes time since the introduction of the virus measured against March 31 2020, the rate of its expansion between week one and week two of the pandemic in the respective countries as well as the disease profile of each nation. We have modeled the ‘early expansion factor’ using air traffic as its instrument because the early stage of the epidemic is presumed to be dominated by imported cases rather than due to community infection. Wooldridge’s (1995) robust score test for endogeneity has confirmed our expectation that the rate of early expansion must be treated as endogenous.

We have also included in our model household size and age structure because both can influence social distancing behavior and practices and could play a mediating role once a community infection is in place. Urbanization and living standard can be expected to further serve as fertile grounds for spreading viruses widely as they could facilitate the interaction and flow of people across a wider environment. Likewise, countries that adhere to international health regulations and provide better access to quality health care are likely to detect and report cases better.

Taking these hypothesized relationships in to account, we have captured the following covariates in our model: time since first reported case, rate of expansion of the disease between week one and week two of the epidemic, urbanicity, socio-demographic index (as a measure of living standard), average household size, the age structure of the population, health care access and quality index, adherence to international health regulations, and prevalence of HIV and asthma.

The data on prevalence of HIV and asthma, as well as on socio-demographic index and the index of health care access and quality, were from Institute of Health Metrics and Evaluation (IHME). (21-23) The data on urbanization were obtained from the Population Reference Bureau, (24) while average household size, age profile, and data on the total population used for the calculation of rates were all from the UN Population Division. (25) The index of adherence to international health regulations was accessed from the World Health Organization (WHO), while the number of confirmed cases was from the John Hopkins database of COVID-19. (26-27)

The 11 variables selected for analyses were chosen out of an initial set of over 20 variables that included the proportion of households with at least one member aged 60 years or over, the proportion of households with at least one member aged 65 years or over, the World Bank’s governance index, as well as the prevalence of TB, diabetes and malaria. STATA’s Lasso software for model selection was used to narrow down the list of covariates. (28) Furthermore, out of the global database covering the 193 countries that we captured as an input in our analyses, for a few countries, data were unavailable for selected variables, namely air traffic (25 countries), adherence to international health regulation (20 countries), and household size (24 countries). To bridge the gap, we performed multiple imputation procedures, particularly by running 1000 imputations and 99 000 iterations using STATA’s multiple imputation routine. (29-30) The resulting median values from the imputation exercise were subsequently used to estimate the prediction model specified earlier. We transformed all the regressors as well as the dependent variables to a log-scale before they were put into the model.

The goodness-of-fit for the predictive model was evaluated using k-fold cross-validation technique. (31-32) The advantage of a k-fold cross-validation technique over other statistical validation procedures is that all observations are used for both training and validation, and each observation is used for validation exactly once. (31) When summarized, our prediction results can be viewed as a function of time since the introduction of the virus, once those other time-invariant covariates are controlled for. Higher order relationships with time are not considered here because the pandemic in the Africa region is in early and expansionary stage. As more and more countries in the world manage to ‘flatten the curve’ the trends and lessons from those experiences can be integrated in covariate-based models like ours.

Estimation of COVID-19 deaths was made using an approach that loosely related to indirect standardization technique, with additional adjustments for differences in health system readiness and living standards. In the absence of age-specific infection data the proposed approach can be expressed in a general form as follows (including in instances where age is treated as a continuous function):

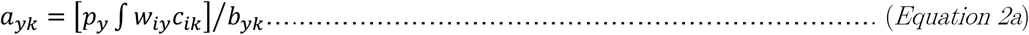

Where,

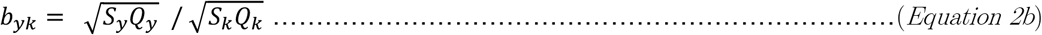

Here, *a*_*yk*_ is the estimated number of deaths for country y (y= 1, 2…47) using country k’s (China and Italy) COVID-19 fatality rate, general health system readiness and socio-economic standards as a reference. *c*_*ik*_ is the standard or reference age-specific fatality rate for age group i in standard country k, in our case obtained from China and Italy. *w*_*iy*_ is the population at age i in country y; *p*_*y*_ is population level COVID-19 prevalence rate and is an estimate we generated using the instrumental variable regression expressed in equation 1. The quantity in the square bracket represents age-standardized estimated number of deaths in country y, if age-specific fatality rates were to follow those of the standard populations. However, our study population differs from the two countries not just with respect to age structure but also living standard and health system readiness. Hence, to adjust the estimated deaths further we developed a correction factor that links the socio-economic status and health system readiness of each country with those two populations, which is represented by *b*_*yk*_. In this case, *b*_*yk*_ is a geometric-mean of the socio-demographic index (S) and the health access and quality index (Q) relative to that of country k. The relevant data for each country was sourced from IHME. (21-22) Analysis was performed using STATA version 16.0.(33)

## III. RESULTS

Figure A1 in the Appendix shows a graphical comparison of observed and predicted confirmed COVID-19 cases (on the log scale). Table A1 presents results from the cross-fold robustness test for the predictive model. The statistical diagnostic tests as well the patterns from the graphical analysis suggest that the predicative model sufficiently captures the observed data. As can be seen from the table, not only are the RMSE, the MAE and the Pseudo-R^2^ consistent across samples, the error margins are also small. Hence, we have used the resulting model to forecast cumulative COVID-19 cases, estimate new infections and deaths from COVID-19 for each of the 47 African countries which have reported at least one case as of March 31, 2020.

### Cumulative COVID-19 cases and point prevalence rates

Table 1 presents cumulative COVID-19 cases and prevalence rates for Africa and its five sub-regions for the period April 1 to June 30 2020. By the end of April cumulated infection across the region will reach 2.5 million cases and will further increase to 16.2 million by June 30. During the same period, cumulative cases in the hardest hit sub-region in the African continent, Northern Africa, is predicted to increase from 1.3 million at the end of April to over 7.5 million by June 30. This is followed by Southern, Western, and Central Africa sub-regions. Throughout the forecast period, Eastern Africa is predicted to experience the lowest case load and prevalence rate in the continent throughout the forecast period. Note these estimates are highly uncertain; the 95% confidence intervals (CI) are shown in Table 1 below.

**Table 1:**
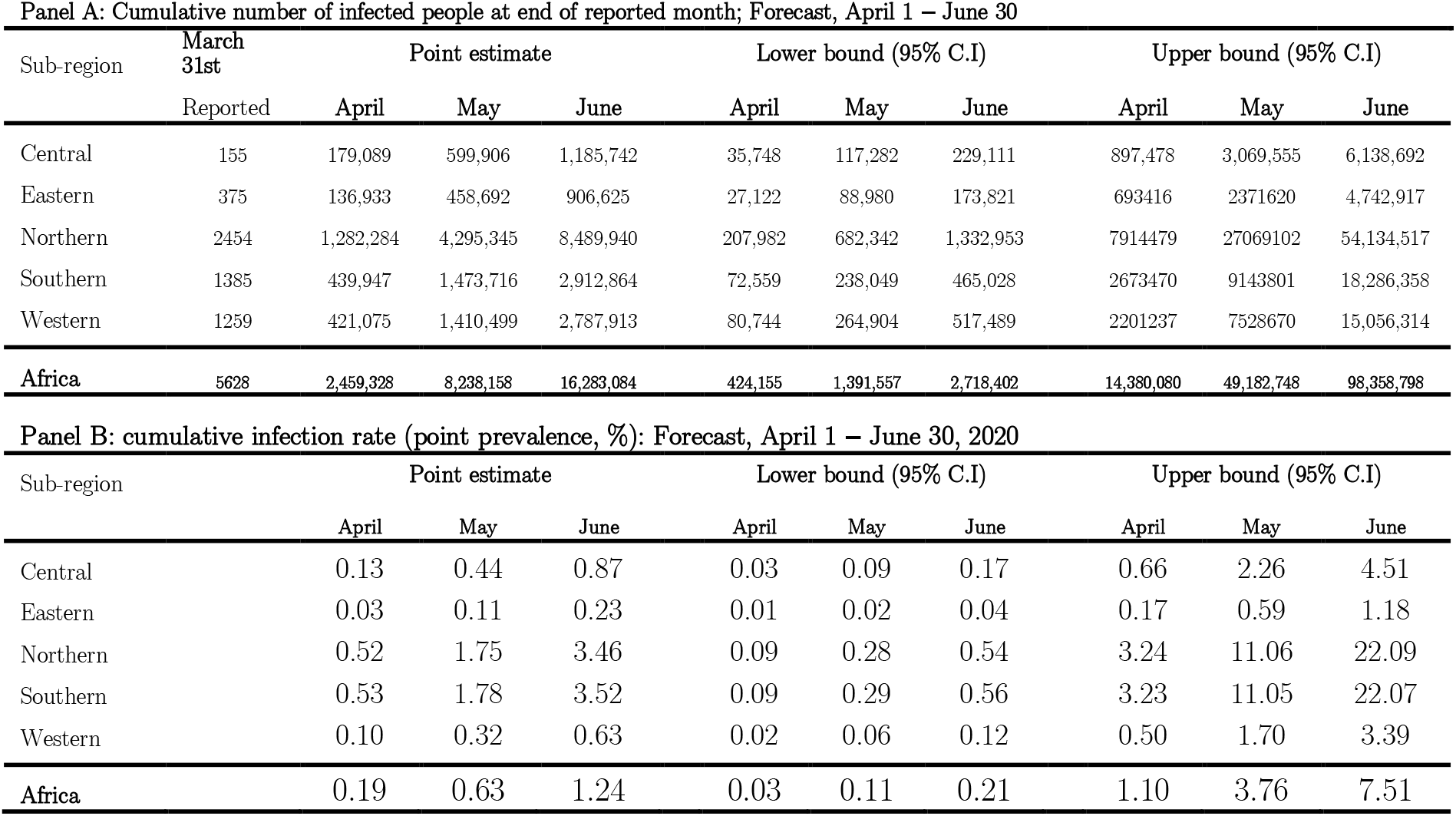
Cumulative cases and prevalence rates for COVID-19, Africa: April 1 – June 30, 2020.

**Table 2:** Number of new infections and incidence rates for COVID-19, Africa: April 1 – June 30, 2020.

| **Panel A: Number of new infection at end of reported month; Forecast, April 1 – June 30** |  |  |  |  |  |  |  |  |  |
| --- | --- | --- | --- | --- | --- | --- | --- | --- | --- |
| Sub-region | Point estimate |  |  | Lower bound (95% C.I) |  |  | Upper bound (95% C.I) |  |  |
|  | April | May | June | April | May | June | April | May | June |
| Central | 178,934 | 420,817 | 585,835 | 35,593 | 81,534 | 111,829 | 897,323 | 2,172,077 | 3,069,137 |
| Eastern | 136,558 | 321,759 | 447,933 | 26,747 | 61,858 | 84,842 | 693,041 | 1,678,204 | 2,371,297 |
| Northern | 1,279,830 | 3,013,060 | 4,194,595 | 205,528 | 474,360 | 650,611 | 7,912,025 | 19,154,623 | 27,065,415 |
| Southern | 438,562 | 1,033,769 | 1,439,149 | 71,174 | 165,490 | 226,979 | 2,672,085 | 6,470,331 | 9,142,557 |
| Western | 419,816 | 989,424 | 1,377,415 | 79,485 | 184,160 | 252,585 | 2,199,978 | 5,327,433 | 7,527,645 |
| **Africa** | **2,453,700** | **5,778,830** | **8,044,927** | **418,527** | **967,402** | **1,326,846** | **14,374,452** | **34,802,668** | **49,176,051** |

| **Panel B: New infection in each month (incidence rate, %): Forecast, April 1 – June 30, 2020** |  |  |  |  |  |  |  |  |  |
| --- | --- | --- | --- | --- | --- | --- | --- | --- | --- |
| Sub-region | Point estimate |  |  | Lower bound (95% C.I) |  |  | Upper bound (95% C.I) |  |  |
|  | April | May | June | April | May | June | April | May | June |
| Central | 0.13 | 0.31 | 0.43 | 0.03 | 0.06 | 0.08 | 0.66 | 1.60 | 2.25 |
| Eastern | 0.03 | 0.08 | 0.11 | 0.01 | 0.02 | 0.02 | 0.17 | 0.42 | 0.59 |
| Northern | 0.52 | 1.23 | 1.71 | 0.08 | 0.19 | 0.27 | 3.24 | 7.83 | 11.04 |
| Southern | 0.53 | 1.25 | 1.74 | 0.09 | 0.20 | 0.27 | 3.23 | 7.82 | 11.03 |
| Western | 0.10 | 0.22 | 0.31 | 0.02 | 0.04 | 0.06 | 0.50 | 1.20 | 1.70 |
| **Africa** | **0.19** | **0.44** | **0.61** | **0.03** | **0.07** | **0.10** | **1.10** | **2.66** | **3.76** |

Figure 1A shows spatial patterns of cumulative COVID-19 cases for the African continent. In Northern Africa, the leading contributor to the burden of COVID-19 is Morocco. By the end of June, Morocco will have 4.5 million cumulative COVID-19 cases, and this is almost double the estimated number for Algeria, a country with the next highest burden, 2.8 by the end of June. In Southern Africa, South Africa and Swaziland are on the lead. By the end of June, these two countries are expected to have respectively around 2.6 million and 250 thousand cumulative COVID-19 cases. In the Western Africa sub-region, cumulative cases will be dominated by Cote d’Ivoire and Ghana, despite Nigeria having a larger population than both countries combined.

**Figure 1.**
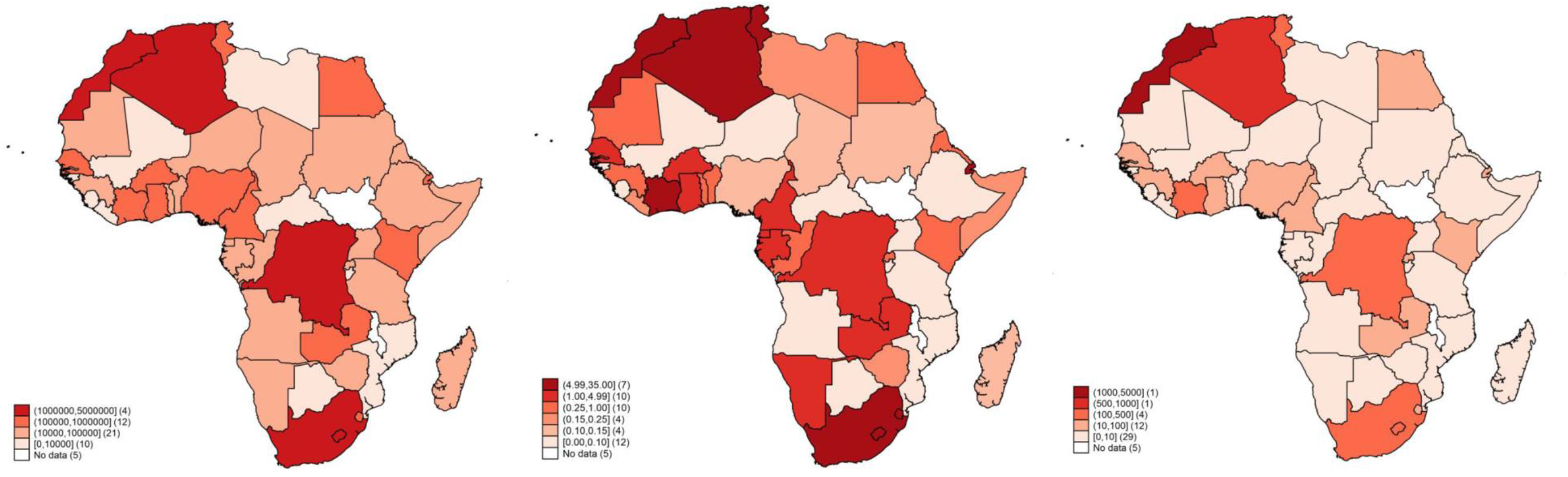
(a) Cumulated COVID cases June 30, 2020; (b) COVID-19 incidence per 1000 population April 2020, and (c) Number of COVID-19 deaths April 2020.

To remove the effect of size, we also estimated prevalence rates shown in Panel B of Table 1 by combining the forecasts on cumulative cases with population estimates which we generated for each month from the UN annual population estimates. This suggests that population level prevalence of COVID-19 in Africa is expected to remain under 1.5 percent (CI 95% 0.03 to 7.6) throughout the prediction period, but there are wide inter-regional differences. In the Northern and Southern Africa sub-regions, cumulative infection rates are expected to be slightly over 3 percent, with the rate in the respective subregions expected to be around 3.5 percent (CI 95% 0.09 to 22.1) as of June 30. This means that even under a very high infection scenario, as observed in the 95% upper confidence interval for the two sub-regions, infection rates are unlikely to reach more than a quarter of the continent’s population.

### New infections and COVID-19 incidence rates across Africa

Across the months, the number of new COVID-19 infections are expected to increase from 2.5 million cases in April, to 5.8 million cases in May, to 8 million cases by the end of June. This represents a 135% increase from April to May and a 39% increase from May to June, which suggests a possible slowing down of the pandemic over time.

Sub-regional and country level differences are expected to deepen as the pandemic becomes more established in the region. In the coming three months, incidence is expected to increase faster in Southern and Northern Africa, followed by the Central Africa sub-region. In each sub-region, we expect some countries to be affected more than others and become hotbeds of new infections (see Figure 1B). In places like Djibouti, the rate of new infections is expected to reach as high as 32.8 (CI 95% of 6.25 to 171.77) cases per 1000 population, followed by Swaziland with a rate of new infection of 26.8 (CI 95% of 4.97 to 144.39), Morocco with 11.97 (CI 95% of 1.98 to 71.59), Algeria with 9.80 (CI 95% of 1.57 to 60.72) and Cote d’Ivorie with 6.65 (CI 95% of 1.07 to 29.68). See Figure 1B.

### Estimated COVID-19 mortality

Deaths for COVID-19 are estimated for each country, using the approach described in Section 2. As can be seen in Figure 2, the adjustment shows relatively higher expected mortality using Italy as a standard compared to China, but generally are closer to each other. Hence, our final country level estimates presented in Figure 1C and the regional summaries in Table 3, were generated using the arithmetic mean of death numbers generated using the two standard populations.

**Table 3:**
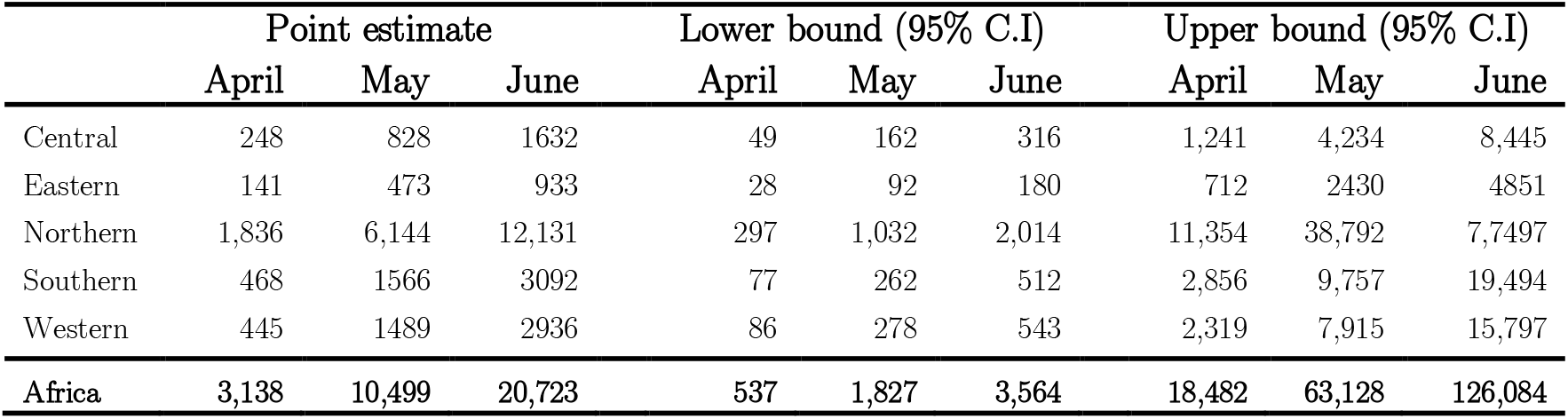
Estimated number of COVID-19 deaths, April- June 2020

**Figure 2:**
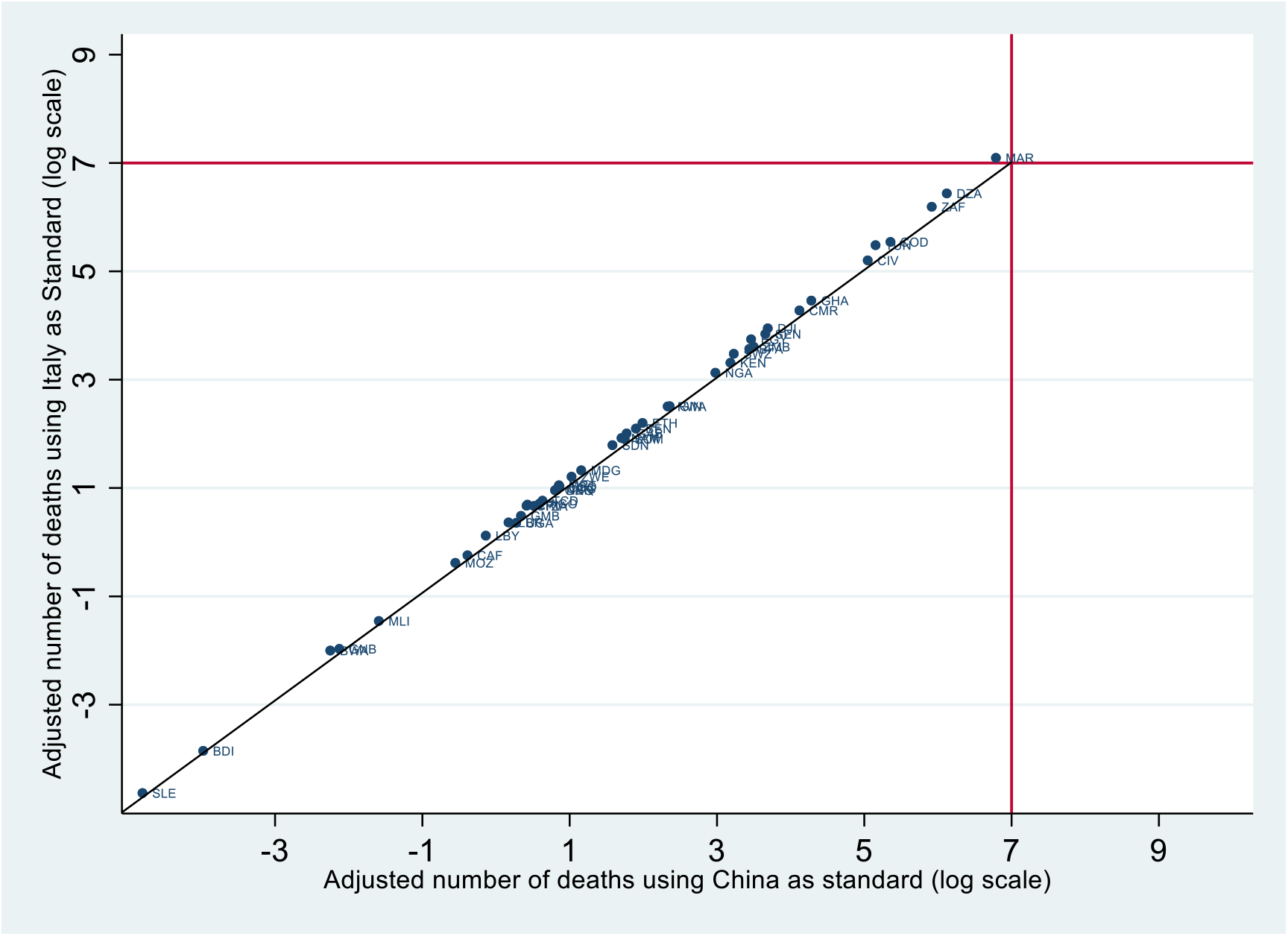
Comparison of estimated number of deaths using China and Italy as standard, April 2020.

The first COVID-19 case in Africa was reported on February 14, 2020 (2, 3). Two months after the first COVID-19 cases were reported in the region, the number of deaths at the end of April is expected to reach over 3,000 deaths (CI 95% of 537 to 18,482). Most deaths will be recorded in the Northern Africa sub-region, which, as reported earlier, also had the largest number of COVID-19 cases and the highest prevalence and incidence rates of all sub-regions in the content. Southern and Western Africa are expected to experience a comparable death toll, while deaths in the Eastern Africa sub-region are expected to be the lowest of the sub-regions in Africa. Some countries such as Algeria, Morocco and South Africa are expected to experience the highest number of casualties since the beginning of the pandemic, in excess of 400 deaths. Deaths in the three countries will comprise of almost two-thirds of all expected deaths in the region. In others, such as the Democratic Republic of Congo, Tunisia and Cote d I’vorie, the death toll will reach between 150 and 300, and contribute 20% to the regional total, while half of the continent (about 29 countries) will experience less than 10 deaths each. The same trend continues in May and June, but with a much higher number of expected deaths.

## IV. DISCUSSION

The COVID-19 pandemic has been spreading rapidly across different parts of the world and the African continent has not been spared. Given the global trajectory, what lies ahead in terms of the course and magnitude of infection in Africa in the days and months to come remains speculative. While there have been attempts elsewhere in capturing the trajectory of the COVID-19 pandemic using agent-based, or mathematical, and or statistical models, this has not been the case for the African region as whole. (4-6, 8) To the best of our knowledge, no study, using a robust methodology, provides estimates on future trend of COVID-19 for the entire region or accounts for its local context.

Modeling and predicting the epidemiology and trajectory of a disease such as COVID-19 is a challenging exercise. Firstly, COVID-19 is a newly identified pathogen and knowledge about the characteristics of the virus, including how the disease is spread and the time from exposure to onset of symptoms are still evolving. Secondly, at the time of writing, less than a handful of countries had reached a plateau in their infection rate, and this makes modeling future trends an open-ended exercise. (1) The fact that some people only experience mild symptoms and that even the best health system can only detect and treat those presenting to facilities also means that the available data on ‘confirmed’ cases represents only a fraction of the true picture of the pandemic.

Thirdly, literature on the association between disease prevalence and population level characteristics that are known to be the staple of social and descriptive epidemiology, and the backbone of predictive modeling exercises are yet to emerge for COVID-19. (4-6, 8) Further, at the individual level, knowledge on the determinants of COVID-19 morbidity and mortality is limited to selected characteristics such as age and pre-existing conditions, and even then, they are drawn from scanty data. The association with community and population level characteristics is even more limited, hence largely speculative. In this context, the task of developing a multi-country covariate-based predictive model for COVID-19 in any country or region of the world, let alone Africa, is likely to be guided by what is feasible, and to the extent, the available data permits it. It also means that there is a need for constant revision of predictive models as new data becomes available. In our case, the work is further complicated because in Africa data on key covariates are either lacking or when they exist, they tend to be biased or derived from other global covariate-based modeling exercises. (21-22)

Despite this, we have made efforts to mitigate these limitations by curating data from various credible sources around the world and assessing them for consistency. In developing our model, we have used data sources from 193 countries globally and used statistical relationships to fill data gaps on covariates in the Africa region. Additionally, our model performance was assessed through rigorous out of sample calibration using k-fold cross-validation techniques and obtaining robust results. We have also restricted our forecast to the first few months (April-June) and refrained from a long-term projection exercise. Firstly, this is because we anticipate new and additional data to emerge in the short term that will lead to better and improved estimates. Secondly, where data is sparse, any long-term projection is more likely to be detached from reality and can easily become a wild guess and is therefore less useful for informing policy actions. Thirdly, if the countries in the region will not be able to put effective strategies soon (or in those who have already initiated them render to be ineffective), the extent and consequence of COVID-19 on the continent would be greater than any predictive model could anticipate.

Within these caveats, our study substantially contributes to the growing literature on understanding the trajectory of the COVID-19 pandemic. This perspective is particularly important for Africa at a time of controlling a pandemic given that Africa the region is home to over 1.3 billion people. (34) In addition, as the COVID-19 pandemic is predicted to start to taper off in the northern hemisphere there are increasing concerns and attention towards Africa and elsewhere in the southern hemisphere that had late introduction and slow uptick of cases. Our model projection is therefore necessary, and timely to guide this attention.

Our results show a heterogeneous picture of projected new infections, cumulative cases, and deaths across the African continent. Our Among our key findings are that African countries that are relatively more urbanized and have a higher socio-demographic index (SDI) are projected to experience a faster growth of the epidemic, at least during the initial periods. To illustrate this trend, Algeria, Morocco, and Tunisia, in the Northern African sub-region, which have a predominantly urban population, and are in close proximity to the high burden European countries, are projected to experience a higher incidence of COVID-19 infections in comparison to their neighbors. In the Southern Africa sub-region, South Africa, which has a proportionately higher urban population and greater international connectivity, is also projected to lead the rest of the pack in terms of new infections rates. In the Eastern Africa sub-region, Kenya, which is relatively more urbanized has a higher per capita incidence rate in comparison to other countries within the sub-region. However, it is noted that some of these countries also have higher living standards and better health system than most countries in the region. Hence the high prevalence rates and estimated deaths in these countries may in part reflect their greater ability to detect and confirm COVID-19 cases.

On the other hand, countries such as Angola, Botswana and Mozambique with a lower international connectivity are projected to have lower rates of new infections during the initial periods. Similar trends are observed in Burundi and Tanzania in the Eastern Africa region. However, as the epidemic grows many of these countries are projected to experience increases in cumulative infections that are likely to overwhelm their health system. Overall, these trends are consistent with other regions of the world, where since the onset of this pandemic, it has been clear that higher connectivity, particularly through air traffic, and higher population densities are key drivers in the introduction and growth of the epidemic. (35-39)

The impact of population density is clearly illustrated by the projected new infections of COVID-19 in places like Djibouti and Rwanda. To compound this, close to 90% of urban settlements in Africa are comprised of informal settlements, most of which are overcrowded and lack basic amenities such as clean water and sanitation which are critical prevention measures for COVID-19. (40-42) Moreover, 60% of jobs in urban areas are in the informal economy, (43) and if African countries were to implement stringent lockdowns these livelihoods would be largely affected and could lead to significant socioeconomic consequences. For instance, those that lose their jobs are likely to move back to their homes in rural areas, where many of the elderly population reside, hence putting them at risk. Similarly, as the economy slows down in several countries due to the COVID-19 pandemic, some sections of migrant workers from African countries might be forced to return to their home countries and hence further complicating the dynamics of the pandemic.

Therefore, large scale measures that are aimed at limiting population movement, across countries as well as increasing the social distance among populations, are not enough, or at best may prove to be impractical to address the pandemic in the African context. Comprehensive response measures should be contextualized and seek to address some of the underlying individual and structural factors that are likely to complicate the epidemic within these environments. It is crucial to balance interventions geared towards preventing the spread of the epidemic with the need to maintaining livelihoods and social cohesion. Measures such as appropriate messaging, provision of adequate water and sanitation subsidies, food as well as targeted restriction of movement (e.g. from urban to rural) would go a long way to mitigate the spread.

Still, even with all these NPI measures, several countries with weak healthcare delivery systems are expected to have a significant number of infections and fatalities. Therefore, the already overstretched healthcare systems in Africa need to anticipate and prepare to handle an increased number of patients as a consequence of the pandemic, on top of the other common disease conditions that are prevalent in the region. Unfortunately, this is a lesson Africa has already had to learn from the 2014 Ebola outbreak in West Africa. It is estimated there were an additional 11,000 deaths from malaria, HIV/AIDS and TB across Sierra-Leone, Guinea and Liberia, (44) a 60% decrease in the number of children treated for diarrhea and acute respiratory infections (45) and a 38% increase in maternal mortality in Guinea and a 111% increase in Liberia. (46-47)

Understanding the factors that accelerate and those that mitigate the spread and mortality related to COVID-19, while accounting for local realities, is fundamental for sound public health measures to tackle the pandemic. In fact, lessons from HIV/AIDS programming have taught us that highly effective health interventions fail if the local context is not recognized. Therefore, decision makers in Africa must realize that one size does not fit all and could lead to disastrous outcomes. They should also find ways and means to maintain the progress made toward universal childhood immunization and the HIV epidemics in the region.

Effective detection and response to epidemics is premised on decision makers having the right epidemiological information. Unfortunately, many countries in Africa have weak health information systems that are not able to collect and process data rapidly to facilitate timely and targeted action. For example, Lesotho, despite being in the same region and with closer proximity to South Africa and Swaziland, countries that are at the epicentre of the COVID-19 pandemic in the Southern Africa sub-region, is yet to register a confirmed case. This is a clear example of the gap in diagnostic capacity and health information system rather than the absence of infection in the country. Hence, the fight to stop the pandemic in Africa should be focused on both strengthening clinical capacity as well reporting systems within the health care system. Innovations around collecting data through digital platforms make it easier to combine different data streams, thereby helping decision-makers gain a better understanding of epidemics and the impact of the interventions. (48-52) With close to 747 million SIM connections in Sub-Saharan Africa, representing 74% of the population on the continent, (53) the response to the COVID-19 pandemic provides an opportunity for countries to retool their data collection systems to meet their surveillance needs.

In addition, the mobile digital technologies could enable confidential self-reporting of symptoms to healthcare providers’, catalyzing further investigation and epidemic control measures before it spreads beyond defined population subgroups. As an information and health communication tool, mobile platforms could also be leveraged to deliver appropriate health messages to the population, relieving direct consultation pressure from health system providers.

Efficient resource allocation and use is only feasible where there is improved visibility across the various components of the health system through high quality health information systems. (54) Ideally, this should cover key areas such as human resources for health, infrastructure and equipment, medicines and other health commodities, and financial resources available. This is particularly needed in epidemic situations, where there are often huge distortions in demand for resources, that if not well addressed could lead to wastage and suboptimal population health outcomes.

Lastly, but equally important in the African context, the global health community and governments addressing the COVID-19 pandemic should be inclusive in their efforts because the efficacy of a response to any emergency is only as good as its weakest link. It is therefore important that governments include marginalized communities, such as refugees, migrants and internally displaced populations (IDPs) in their policies and actions. Across Africa, approximately 12.3 million people remain forcibly displaced, including 8.1 million internally displaced people and 4.2 million refugees. (55) Moreover, Sub-Saharan

Africa alone hosts more than 26% of the world’s refugee population. (56-57) These populations are likely to face a different set of constraints that restricts their ability to prevent themselves from getting infected and accessing health care. The fact that they also live in under difficult circumstances means that governments across the region should maintain critical supply corridors for humanitarian assistance and address the needs of the most vulnerable within their borders. In the longer term this pandemic should also be a call for African governments to further strengthen engagement with international humanitarian actors in order to foster strong partnership building on local capacities. Within this supranational context, the issue of ‘livelihood corridor’ also needs to be looked at carefully and more broadly, given that AfCFTA (the African Continental Free Trade Agreement) which is expected to govern trade and flow of goods and services in the region will come into effect on July 1 2020 (African Union 2020). Hence, current and planned NPIs in the region need to consider such regional initiatives to ensure that responses to the pandemic at country and regional level are coherent and able to balance saving the lives and livelihood of the people of the African continent.

## V. CONCLUSIONS

It is true that some of the most successful responses to global health threats including recent ones like Ebola and HIV/AIDS have been characterized by multi-stakeholder partnerships. African health systems have been in the forefront in the implementation of such programs and would be best served to leverage that experience and resources in responding to COVID-19. Bringing all stakeholders together to ensure effective coordination, pooling of resources and delivery of evidence-based interventions is imperative for any sustainable response to such pandemic situations.

Within, the African context, policy makers should also consider (a) impact of strategies that could potentially deepen health inequalities and (b) continuously use data driven approaches to identifying their unique most at risk/vulnerable groups e.g. IDP, HIV positive, food insecure to support them in an equitable manner and bring them into the COVID-19 prevention framework. At a time of a pandemic no one community can be marginalized.

Finally, in as much as health systems are dealing with an emerging situation, measurement is a fundamental tool to guide strategic response actions. The measurement framework should be agile yet comprehensive, rather than being piecemeal. It should seek to combine data from multiple sources within the healthcare system into a joint assessment framework such that a consistent and informative narrative emerges to tackle the epidemic. The age-old call for accurate health metrics to address population health still remains valid. Our best estimates presented in the paper are a forecast, and their ability to capture reality is as good as the underlying data. It is thus imperative to review the model and the results as new data becomes available and update the estimates accordingly.

## Data Availability

All data is publically available

http://ghdx.healthdata.org/

https://www.prb.org/data/

https://www.un.org

https://coronavirus.jhu.edu/data

https://www.who.int/healthinfo/statistics/en/

## AUTHORS CONTRIBUTIONS

YK conceptualised the study, designed methods and approach; guided the statistical analysis and drafted the manuscript. UA synthesised the literature, contributed to conceptualisation and drafted the manuscript. TA accessed data, contributed to methods and design, undertook the statistical analysis and drafted the manuscript. LW, TG, FS, AL SL, and RGW reviewed the draft and contributed to the scientific content of the manuscript. All authors contributed to the discussion, read and approved the final draft.

### ACKNOWLEDGMENTS

This study was finalised when YK was in a government designated quarantine centre for COVID-19 at Sydney InterContinental Hotel, upon his return to Australia. YK would like to acknowledge the Commonwealth Government of Australia and the New South Wales Government for supporting his stay at the hotel free of charge and creating an environment that enabled him to continue with his research and teaching activities, while doing their part to combat COVID-19.

## PATIENT AND PUBLIC INVOLVMENT STATEMENT

Patients or the public WERE NOT involved in the design, or conduct, or reporting, or dissemination plans of our research.

## ETHICAL APPROVAL

Analyses was undertake using publicly available secondary data, hence no ethics approval was sought.

---

**Appendix Figure A1:**
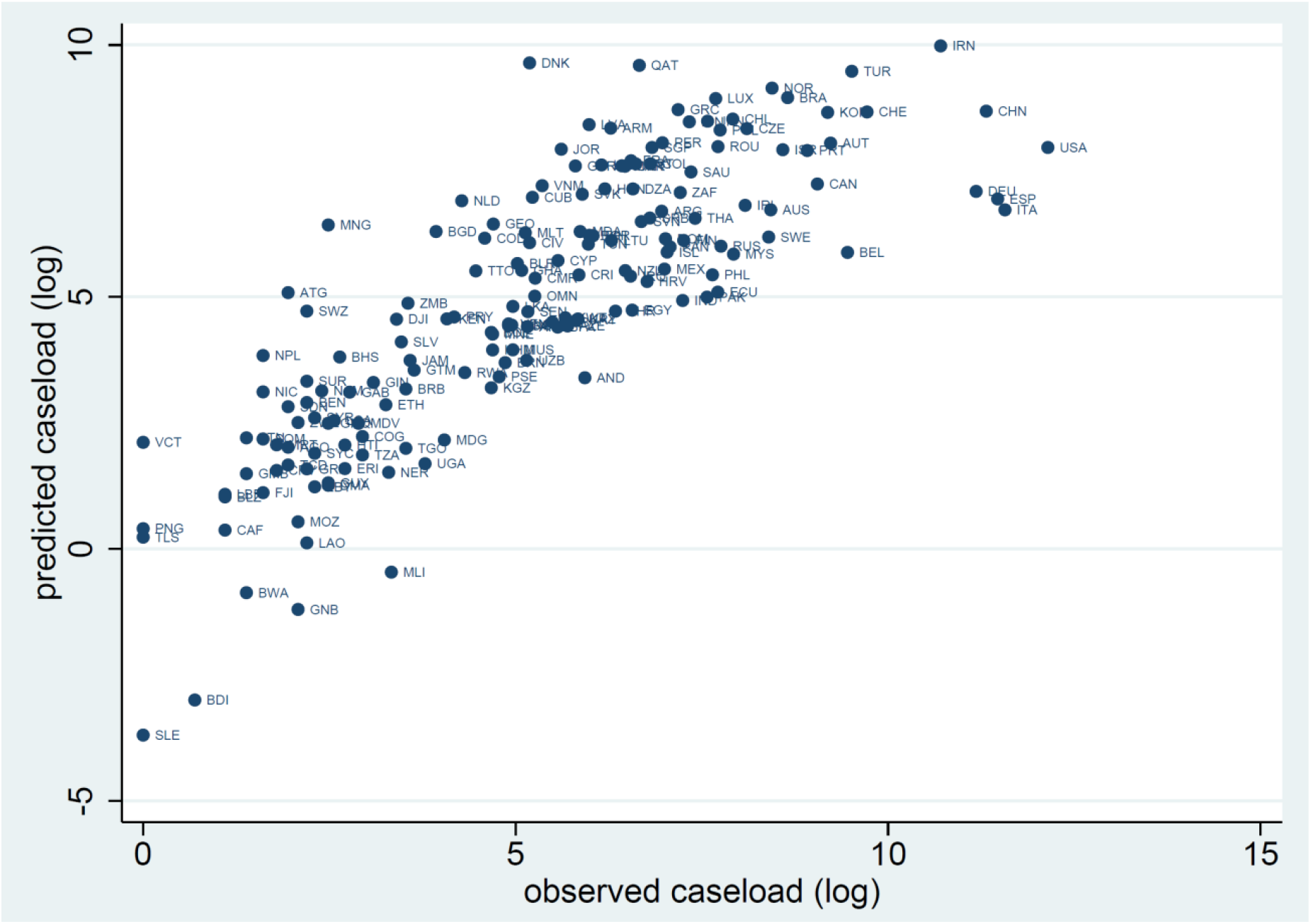
Observed and predicted COVID-19 cases, March 31, 2020.

**Appendix Table A1:** Predication model performance from K-fold cross-validation test

| Robustness<br>parameter | Sub-sample number |  |  |  |  |  |  |  |  |
| --- | --- | --- | --- | --- | --- | --- | --- | --- | --- |
|  | 1 | 2 | 3 | 4 | 5 | 6 | 7 | 8 | 9 |
| RMSE | 1.476 | 1.457 | 1.483 | 1.449 | 1.483 | 1.484 | 1.485 | 1.487 | 1.485 |
| MAE | 1.114 | 1.141 | 1.128 | 1.137 | 1.159 | 1.139 | 1.146 | 1.134 | 1.143 |
| Pseudo-R<sup>2</sup> | 0.718 | 0.721 | 0.717 | 0.719 | 0.730 | 0.719 | 0.728 | 0.728 | 0.726 |

Note : RMSE, Root Mean Square Error; MAE, Mean Absolute Error.

## REFERENCES

1. WHO WHO COVID-19 data accessed April 3 2020

2. African CDC https://africacdc.org/covid-19/ accessed April 2 2020

3. WHO EMRO http://www.emro.who.int/index.html accessed April 6 2020

4. Ferguson N.M, Laydon. D, Nedjati-Gilani. G, Imai. N, Ainslie. K, Baguelin. M, Bhatia. S, Boonyasiri. A et. al., Impact of non-pharmaceutical interventions (NPIs) to reduce COVID-19 mortality and healthcare demand. London: Imperial College COVID-19 Response Team, March 16 (2020). Imperial College accessed March 23 2020

5. Tian. H, Liu. Y, Wu. C, Chen. B, Kraemer. M et. al, Early evaluation of transmission control measures in response to the 2019 novel coronavirus outbreak in China. https://www.medrxiv.org/content/10.1101/2020.01.30.20019844v4

6. Murray. C. Jl, IHME COVID-19 health service utilization forecasting team. Forecasting COVID-19 impact on hospital bed-days, ICU-days, ventilator-days and deaths by US state in the next 4 months medRxiv 2020.03.27.20043752; https://doi.org/10.1101/2020.03.27.20043752

7. WHO. Coronavirus disease (COVID2019) situation report-30. https://www.who.int/docs/defaultsource/coronaviruses/situation reports/20200219sitrep30-covid19.pdf?sfvrsn=3346b04f_2 accessed March 31 2020

8. Martinez-Alvarez. M, Jarde. A, Usuf. E, Brotherton. H, Bittaye. M, Samateh Al, Antonio. M et.al., COVID-19 pandemic in West Africa Lancet Glob Health 2020, April 1, 2020

9. Anderson RM, Heesterbeek H, Klinkenberg D, Hollingsworth Hollingsworth T. How will country-based mitigation measures influence the course of the COVID-19 epidemic? Lancet. Comment| Volume 395, ISSUE 10228, P931–934, March 21, 2020

10. European Centre for Disease Prevention and Control. Daily risk assessment on COVID19 https://www.ecdc.europa.eu/en/cases-2019-ncov-eueea accessed April 2 2020

11. Zou L Ruan f, Huang M, Liang L, Huang H, Hong Z, Yu J, et.al., SARSCoV2 viral load in upper respiratory specimens of infected patients. N Engl J Med 2020; 382:1177–1179

12. Kinfu. Y, Opiyo. C, & Wamukoya, M. (2014). Child Health and Mortality in sub-Saharan Africa: Trends, causes, and forecasts. In C. O. Odimegwu, & J. Kekovole (Eds.), Continuity and change in sub-saharan African Demography (Vol. 17, pp. 60-77). (Routledge African Studies; Vol. 17). New York, USA: Routledge https://doi.org/10.4324/9781315879444

13. Kastelic. K, Himelein. T, Mauro. T, Abubakarr. T,. The socio-economic impacts of Ebola in Sierra Leone : results from a high frequency cell phone survey (round three) (English). Washington, D.C: World Bank Group. http://documents.worldbank.org/curated/en/873321467999676330/The-socio-economic-impacts-of-Ebola-in-Sierra-Leone-results-from-a-high-frequency-cell-phone-survey-round-three accessed March 22 2020

14. Wooldridge. J 2010 Econometric Analysis of cross section and panel data. 2nd nde. Cambridge, MA: MIT press

15. Cameron A.C and Trivedi P.K. 2010 Micro econometrics using Stata. Rev ed College Station, TX: Stata Press

16. Davidson R and Mackinnon J.G. 1993 Estimation and inference in econometrics New York: Oxford University press

17. Martens. E.P, Pestman. W.R, De Boer. A, Belitser. S.V, & Klungel O.H. (2006). Instrumental Variables: Application and Limitations. Epidemiology, 17(3), 260–267

18. Cameron A.C and Trivedi P.K. 2005 Microeconometrics methods and applications New York: Cambridge University press

19. Greene. WH 2018 Econometric Analysis 8th the. New York: Pearson

20. Global Burden of Disease Collaborative Network. Global Burden of Disease Study 2015 (GBD 2015) Socio-Demographic Index (SDI) 1980–2015. Seattle, United States: Institute for Health Metrics and Evaluation (IHME), 2016.

21. GBD 2016 Healthcare Access and Quality Collaborators. Measuring performance on the Healthcare Access and Quality Index for 195 countries and territories and selected subnational locations: a systematic analysis from the Global Burden of Disease Study 2016. Lancet. 2018 Jun 2;391(10136):2236–2271.

22. Global health data exchange http://ghdx.healthdata.org/ accessed March 24 2020

23. Population reference bureau https://www.prb.org/data/ accessed March 24 2020

24. UN Population division https://www.un.org accessed March 25 2020

25. WHO Health Statistics https://www.who.int/healthinfo/statistics/en/ accessed March 22 2020

26. Johns Hopkins Coronavirus resource center https://coronavirus.jhu.edu/data accessed March 31 2020

27. Lasso Reference Manual, Release 16 College Station, TX: Stata Press.

28. Rubin. D.B Multiple imputation for non-response in surveys 1987. New York: Wiley

29. Schafer. J.L Analysis on incomplete multivariate data. 1997. pnoca Raton RatonF. Chapman &Hall/ CRC

30. Hastie T, Tibshirani R, Friedman J. The elements of statistical learning: data mining, inference, and prediction. 2nd ed. New York, NY: Springer; 2009. 219–260

31. Rao C, Adair T, Kinfu Y. Using historical vital statistics to predict the distribution of under-five mortality by cause. Clin Med Res. 2011 Jun; 9(2):66–74.

32. Stata Corp. 2019. Stata Statistical Software: Release 16. College Station, TX: StataCorp LLC

33. UN Population https://www.un.org/en/sections/issues-depth/population/index.html accessed April 1 2020

34. Chinazzi. M, Davis. JT, Ajelli. M, Gioannini. C, Litvinova. M, Merler. S, pastore. PY, et.al.,The effect of travel restrictions on the spread of the 2019 novel coronavirus (COVID-19) outbreak. Science. 2020 Mar 6

35. Wu. T, Leung. K, Leung. G.M, Now casting and forecasting the potential domestic and international spread of the 2019-nCoV outbreak originating in Wuhan, China: A modeling study. Lancet 395, 689–697 (2020).

36. Du. Z, Wang. L, Cauchemez. S, Xu. X, Wang. X, Cowling. B.J, Meyers. L A, Risk for Transportation of 2019 Novel Coronavirus Disease from Wuhan to Other Cities in China. Emerg Infect Dis. 2020 May 17;26(5).

37. Kucharski AJ, Russell TW, Diamond C, Liu Y, Edmunds J, Funk S, Eggo RM et.al, Early dynamics of transmission and control of COVID-19: a mathematical modelling study. Lancet Infect Dis. 2020 Mar 11.

38. Gilbert. M, Pullano. G, Pinotti. F, Valdano. E, Poletto. C, Boëlle. P, D’Ortenzio. E et.al., Preparedness and vulnerability of African countries against importations of COVID-19: a modelling study Lancet 2020; 395: 871–77

39. Simiyu. S, Cairncross. S & Swilling. M. Understanding Living Conditions and Deprivation in Informal Settlements of Kisumu, Kenya. Urban Forum 30, o223–241 (2019).

40. Wakhungu. J, Huhhins. C.D, Lumumba. J 2010 Approaches to informal urban settlements in Africa: experiences from Kigali and Nairobi. Technical report. African Centre for Technology Studies (ACTS)

41. Güneralp. B, Lwasa. S, Masundire. S, Parnell. S, and Seto. K.c Environmental Research Letters, Volume 13, Number 1 Published 20 December 2017

42. AEO 2016 Africa Economic Outlook: Sustainable Cities and Structural Transformation 389 Report

43. Parpia. As, Ndeffo-Mbah. Ml, Wenzel. NS, Galvani. Ap. Effects of Response to 2014–2015 Ebola Outbreak on Deaths from Malaria, HIV/AIDS, and Tuberculosis, West Africa. Emerg Infect Dis. 2016;22(3):433–441.

44. O’Fallon. B, Barry. MA, Brodish. P, Hazerjian. J. Rapid Assessment of Ebola-Related Implications for Reproductive, Maternal, Newborn and Child Health Service Delivery and Utilization in Guinea. PLoS Curr. 2015 Aug 4;7.

45. Mullan. Z The Cost of Ebola Lancet Global Health. Editorial Volume 3, Issue 8. Aug 2015.

46. Elston. JW, Cartwright. C, Ndumbi. P, Wright. J. The health impact of the 2014–15 Ebola outbreak. Public Health 2017; 143: 60–70.

47. Mekuria. LA, Rinke de Wit. TF, Spieker. N, Koech. R, Nyarango. R, et al., Analyzing data from the digital healthcare exchange platform for surveillance of antibiotic prescriptions in primary care in urban Kenya: A mixed-methods studys. Plos one 14(11) 2019.

48. Guo. X, Chen. S, Zhang. X, Ju. X, Wang. X Exploring Patients’ Intentions for Continuous Usage of mHealth Services: Elaboration-Likelihood Perspective Study JMIR Mhealth Uhealth 2020;8(4):e17258

49. Lee. Jm, Newman. MW, Lee. Jm, Gebremariam. A, Choi. P, Lewis. D, Nordgren. W, Costik. J, et.al., Real-World Use and Self-Reported Health Outcomes of a Patient-Designed Do-it Yourself Mobile Technology System for Diabetes: Lessons for Mobile Health. Diabetes Technol Ther. 2017 Apr;19(4):209–219.

50. Cahn. A, Akiroy. A, Raz. I Digital health technology and diabetes management. J Diabetes. 2018 Jan;10(1) 10–17

51. Achoki T. Health Technology in Emerging Global Epidemics: An African Policy Perspective. Georgetown Journal of International Affairs. Published 2016 July 22. Available at: https://www.georgetownjournalofinternationalaffairs.org/online-edition/health-technology-in-emerging-global-epidemics-an-african-policy-perspective

52. GMSA report https://www.gsma.com/mobileeconomy/sub-saharan-africa/ accessed April 1 2020

53. Achoki. T, Hovels. A, Masiye. F, Lesego. A, Leufkens. H, Kinfu. Y. Technical and scale efficiency in the delivery of child health services in Zambia: results from data envelopment analysis. BMJ Open. 2017;7(1):e012321.

54. UNHCR Global trend https://www.unhcr.org/globaltrends2018/ accessed March 31

55. World Economic Forum https://www.weforum.org/agenda/2019/09/newly-released-data-show-refugee-numbers-at-record-levels/ accessed April 1 2020

56. OCHA Southern Africa Humanitarian Snapshot, accessed March 22 2020.

57. African Union, African Contiental Free Trade Agreement, https://au.int/en/cfta accessed April 5 20202.

